# Evaluation of the accuracy of 99DOTS, a novel cellphone-based strategy for monitoring adherence to tuberculosis medications

**DOI:** 10.1101/19011197

**Authors:** Beena E. Thomas, J. Vignesh Kumar, M. Chiranjeevi, Daksha Shah, Amit Khandewale, Jessica E. Haberer, Kenneth H. Mayer, Ramnath Subbaraman

**Author notes:** **Correspondence:** B.E. Thomas, Department of Social and Behavioural Research, ICMR- National Institute for Research in Tuberculosis, No. 1, Mayor Sathiyamoorthy Road, Chetpet, Chennai – 600 031, India. **Alternate corresponding author:** R. Subbaraman, Department of Public Health and Community Medicine, Tufts University School of Medicine, Boston, MA 02111. These authors contributed equally to this work (Beena E. Thomas, Ramnath Subbaraman).

## Abstract

99DOTS is a cellphone-based strategy for monitoring tuberculosis medication adherence. In a cohort of 650 Indian tuberculosis patients, we compared 99DOTS’ adherence record against results of urine isoniazid tests collected during unannounced home visits. 99DOTS had suboptimal accuracy for measuring adherence, partly due to poor patient engagement with 99DOTS.

## INTRODUCTION

Poor adherence to TB therapy is associated with increased risk of death, disease relapse, and development of drug resistance [1]. Historically, national TB programs, including India’s, have monitored adherence using directly observed therapy (DOT), often with a facility-based approach, in which patients visit health facilities, where healthcare providers (HCPs) watch them take every medication dose [2]. However, DOT raises implementation and ethical challenges for health systems and TB patients in resource-limited settings and may not result in superior outcomes compared to self-administered therapy [2].

Digital adherence technologies (DATs) may facilitate more patient-centered approaches to TB care, by allowing patients to take medications at a place of their choice while having adherence monitored remotely [2,3]. 99DOTS is a cellphone-based DAT that has been used to monitor >150,000 TB patients in India’s government program since 2015, including nearly all people living with HIV (PLHIV) taking TB therapy in the public sector [4]. Dispensing each medication dose reveals a hidden phone number that the patient calls to report pill taking; a computer registers this event. HCPs visualize this adherence record remotely to identify, and intervene with, non-adherent patients. 99DOTS’ advantages include capacity to work with feature (non-smart) phones, relatively low cost, and enabling of remote, real-time monitoring by HCPs [4].

While 99DOTS has potential advantages, its value for monitoring treatment depends on its accuracy for measuring adherence. DATs may over-report adherence (i.e., reporting pills being taken when patients are actually non-adherent) or under-report adherence (i.e., reporting pills being missed when patients are actually adherent) [2]. We present findings of a cohort study conducted in India that assessed 99DOTS’ accuracy by comparing its recorded dosing histories against results of urine isoniazid tests collected during unannounced visits to TB patients’ homes.

## METHODS

Ethics committees at the National Institute for Research in TB (NIRT), Brigham and Women’s Hospital, and Tufts University approved this protocol.

To enroll a geographically diverse cohort including PLHIV, we sequentially recruited presumed drug-susceptible TB patients from 11 TB treatment centers in Mumbai (none of whom were PLHIV) and five HIV treatment centers in Chennai and Vellore (all of whom were PLHIV). Patients were enrolled at different time points in the six-month treatment course, during treatment initiation or medication refill visits. We aimed to achieve roughly equal representation of home visits in the first two months (intensive phase) and last four months (continuation phase) of therapy, since medication adherence and 99DOTS engagement may wane with clinical improvement later in treatment. We included patients without and with a prior TB treatment history, taking Category 1 and 2 TB therapies, respectively.

At enrollment, we collected informed consent for a baseline questionnaire and future unannounced home visit. Patients became eligible for a home visit three weeks after enrollment to minimize short-term changes in 99DOTS engagement (i.e., calling) or adherence from anticipation of the visit (Hawthorne effect). The exact day of the visit was selected using a random number generator. During home visits, field researchers administered a structured questionnaire to assess self-reported 99DOTS engagement, self-reported medication adherence, and reasons for suboptimal 99DOTS engagement and medication adherence.

At the end of the home visit, the patient’s urine sample was collected for testing using the point-of-care IsoScreen test (GFC Diagnostics, UK). Mixing urine with IsoScreen reagents results in a green, blue, or purple color change if isoniazid is in the sample; we classified such patients as being “adherent per urine testing.” Patients with yellow IsoScreen test results were classified as “non-adherent per urine testing.” Isoniazid is detectable in urine in nearly all patients between 6—48 hours after ingestion and undetectable in nearly all patients >72 hours after ingestion [5,6]. However, urine isoniazid test results <6 hours and 48—72 hours after ingestion are variable (the “gray zone”) [5,6]. We therefore excluded from analysis urine test results for patients whose 99DOTS record only reported doses taken within “gray zone” timings, without doses reported 6—48 hours before the home visit. This resulted in exclusion of 8% of urine test results from analysis.

We classified patients whose 99DOTS record reported at least one dose taken 6—48 hours before the home visit as being “adherent per 99DOTS”. Patients whose 99DOTS record reported that the last dose was taken >72 hours before the home visit were classified as “non-adherent per 99DOTS”. In addition to patient-reported doses (via phone calls) 99DOTS allows HCPs to report doses, which they often do after calling patients to check on their adherence. HCP-reported doses are represented using light green in the 99DOTS record compared to dark green for patient-reported doses. We analyzed 99DOTS’ operating characteristics first using only patient-reported doses and subsequently including both patient- and HCP-reported doses.

To determine 99DOTS’ operating characteristics for measuring adherence, we estimated the sensitivity, specificity, positive predictive value, negative predictive value, accuracy (proportion correctly classified), and likelihood ratios of the 99DOTS record compared to urine isoniazid test results as the more rigorous biomarker-based comparator. We stratified findings by subgroups: PLHIV versus non-PLHIV, intensive versus continuation treatment phase, and patients without versus with a prior TB treatment history. We used Chi-squared to test for statistically significant differences in prevalence of medication adherence and 99DOTS operating characteristics between subgroups. We used McNemar’s test to assess for significant differences in 99DOTS accuracy using patient-reported doses alone versus patient- and HCP-reported doses.

**Table.** Prevalence of tuberculosis medication adherence and operating characteristics of 99DOTS for measuring medication adherence

| Sample | Number of patients in sample (N) | Prevalence of adherence by urine isoniazid testing<sup>a</sup> | Prevalence of engagement with 99DOTS<sup>b</sup> | Sensitivity | Specificity | Positive predictive value | Negative predictive value | Accuracy | Positive likelihood ratio | Negative likelihood ratio |
| --- | --- | --- | --- | --- | --- | --- | --- | --- | --- | --- |
|  |  | % (95%CI) | % (95%CI) | % (95%CI) | % (95%CI) | % (95%CI) | % (95%CI) | % (95%CI) | % (95%CI) | % (95%CI) |
| Overall cohort (patient-reported doses only)<sup>c</sup> | 597 | 88 (85—91) | 66 (62—70) | 70 (66—74) | 61 (48—72) | 93 (91—95) | 21 (18—25) | 69 (65—72) | 1.8 (1.3—2.4) | 0.5 (0.4—0.6) |
| Overall cohort (patient- and provider-reported doses) | 597 | 88 (85—91) | 82 (78—85) | 85 (81—88) | 39 (28—52) | 91 (90—93) | 26 (20—33) | 79 (76—82) | 1.4 (1.2—1.7) | 0.4 (0.3—0.6) |
| TB patients living with HIV<sup>c</sup> | 287 | 83 (78—87) | 60 (54—65) | 65 (59—71) | 66 (51—79) | 90 (86—93) | 28 (23—34) | 65 (59—71) | 1.9 (1.3—2.8) | 0.5 (0.4—0.7) |
| TB patients without HIV infection<sup>c</sup> | 310 | 93 (90—96) | 72 (67—77) | 74 (68—79) | 48 (26—70) | 95 (93—97) | 12 (7—18) | 72 (67—77) | 1.4 (0.9—2.1) | 0.6 (0.3—0.9) |
| TB patients in the intensive phase of therapy<sup>c</sup> | 334 | 89 (85—92) | 70 (65—75) | 73 (68—78) | 55 (38—71) | 93 (90—95) | 21 (16—27) | 71 (66—76) | 1.6 (1.1—2.4) | 0.5 (0.3—0.7) |
| TB patients in the continuation phase of therapy<sup>c</sup> | 263 | 87 (83—91) | 61 (55—67) | 65 (59—71) | 67 (48—82) | 93 (89—96) | 22 (17—27) | 65 (59—71) | 2.0 (1.2—3.2) | 0.5 (0.4—0.7) |
| TB patients without prior treatment history<sup>c</sup> | 462 | 89 (86—92) | 66 (61—70) | 69 (64—73) | 61 (46—75) | 94 (91—96) | 19 (15—23) | 68 (64—72) | 1.8 (1.2—2.6) | 0.5 (0.4—0.7) |
| TB patients with a prior TB treatment history<sup>c</sup> | 135 | 84 (76—90) | 67 (59—75) | 73 (63—81) | 59 (36—79) | 90 (84—94) | 30 (21—40) | 70 (62—78) | 1.8 (1.1—3.0) | 0.5 (0.3—0.7) |

<sup>a</sup>Note that this refers to any dose of TB medications taken within 6—48 hours prior to the home visit, given the operating characteristics of the urine isoniazid test.
<sup>b</sup>Engagement with 99DOTS refers to any call made between 6—48 hours prior to the home visit
<sup>c</sup>These operating characteristics were estimated only using patient-reported doses.

## RESULTS

Determination of our sample is reported in the supplementary appendix. We report results for 597 patients. Median age was 35 years (range 18 to 83 years), 346 (58%) were men, 334 (56%) were in the intensive phase, 287 (48%) were PLHIV, and 135 (23%) had a prior TB treatment history. Adherence (positive urine isoniazid test) for the overall sample was 88% (95% CI: 85—91) (Table). Among subgroups, adherence and 99DOTS engagement were significantly lower in PLHIV than non-PLHIV (both p≤0.001). 99DOTS engagement was lower in the continuation phase than the intensive phase (p=0.02).

Findings for sensitivity, specificity, positive predictive value, and negative predictive value of 99DOTS for measuring adherence were 70% (95%CI: 66%—74%), 61% (95%CI: 48%—72%), 93% (95%CI: 91%—95%), and 21% (95%CI: 18%—25%), respectively, using patient-reported doses alone. Using patient- and HCP-reported doses, the sensitivity of 99DOTS increased (p≤0.0001), although its specificity decreased considerably (p≤0.0001). 99DOTS had lower sensitivity in the continuation phase compared to the intensive phase (p=0.048). In PLHIV, 99DOTS had lower sensitivity (p=0.03) and lower positive predictive value (p=0.02) but higher negative predictive value (p=0.004) when compared to non-PLHIV.

## DISCUSSION

In this cohort study, medication adherence was relatively high, including in patients with a prior TB treatment history and in the continuation phase—subgroups for whom adherence is often assumed to be suboptimal. However, this prevalence of adherence may not reflect patients’ adherence throughout treatment, given that we collected a single measurement on each patient using a test that is positive for any dose taken in the prior 48—72 hours. Studies also suggest that risk of poor TB treatment outcomes increases even with mild non-adherence (e.g., ≥10% of doses missed) [1]. Our findings therefore highlight a need to further improve adherence in this population, especially for PLHIV.

Patient engagement with 99DOTS was lower than the urine-based measure of adherence—reflected in 99DOTS’s suboptimal sensitivity—particularly for PLHIV and in the continuation phase. This raises considerable challenges for HCPs in using 99DOTS to identify and address non-adherence. 99DOTS’ negative predictive value suggests that HCPs have to reach out to about five patients reported as being non-adherent by 99DOTS to identify one patient who is not taking medications, because most patients who did not call 99DOTS were actually taking medications. Prior studies have found that suboptimal engagement with DATs is context-dependent. For example, TB patients monitored using a two-way SMS strategy in Pakistan [7] and men who have sex with men taking HIV preexposure prophylaxis monitored using electronic pillboxes in the U.S. [8] both under-reported adherence via these technologies for context-specific reasons.

The specificity indicates suboptimal benefits of 99DOTS for identifying and intervening with non-adherent patients. When only patient-reported doses were included, 99DOTS missed identifying about four out of ten patients who were not taking TB medications, presumably because these patients called 99DOTS without ingesting doses. While 99DOTS’ other operating characteristics improved when HCP-reported doses were included, the specificity decreased further, such that 99DOTS missed identifying about six out of ten patients who were not taking medications—a finding that may represent social desirability by patients against admitted to non-adherence when HCPs contacted them. The positive and negative likelihood ratios suggest that the 99DOTS record provides only modest (15% or less) changes in the post-test probabilities of adherence and non-adherence, respectively.

Future analyses should explore patient characteristics associated with higher adherence and 99DOTS engagement. 99DOTS’ suboptimal accuracy in the current study is limited to a single measurement and may not reflect effects that 99DOTS and associated HCP follow-up may have on adherence or treatment success. Rigorous trials are needed to assess 99DOTS’ impact on these outcomes.

In summary, we found suboptimal operating characteristics of 99DOTS for measuring TB medication adherence in a multi-site cohort study. Our study highlights benefits of urine isoniazid testing for understanding TB medication adherence and for assessing the accuracy of DATs, especially given growing interest in these technologies [2,3]. Our findings raise concerns about India’s large-scale 99DOTS deployment, especially given a lack of high-quality data regarding its impact on TB treatment outcomes. Our findings also highlight an urgent need to strengthen 99DOTS’ implementation, with ongoing assessment to evaluate whether its accuracy—and value for monitoring adherence—can be improved.

## Data Availability

Requests for access to the de-identified dataset can be made by contacting Dr. Beena Thomas, although access to these data may be limited by NIRT/Indian Council of Medical Research policies.

## Acknowledgments

We thank all the participants who have consented to take part in this study and offered their support in the conduct of the study. We acknowledge the immense contribution of all the field investigators from the two study sites for their efforts in collecting data and conducting follow-up visits for all study participants. The Mumbai team included Gunjan Rahul Gaurkhede, Apurva Shashikant Walgude, Yogesh Nivrutti Salve, Jagannath Dattatraja Kumbar, Kamble Rakesh Suresh, Shweta Shantaram Bagade, and Meena Atul Kamble. The Tamil Nadu team included J. Hephzibah Mercy, C. Jeganathan, S. Yokeshwaran, S. Kokila, K. Sathiyamoorthy, E. Michael Raj, P. Brindadevi, K. Jegan, B.Sathyan Raj Kumar, Mariyamma Paul, and Ranjith Kumar. We greatly appreciate statistical advice provided by Professor Misha Eliasziw, Tufts University School of Medicine.

